# Newborn DNA methylation age differentiates long-term weight trajectory: The Boston Birth Cohort

**DOI:** 10.1101/2023.11.02.23297965

**Authors:** Anat Yaskolka Meir, Guoying Wang, Xiumei Hong, Xiaobin Wang, Liming Liang

## Abstract

**Background:** Gestational age (GEAA) estimated by newborn DNA methylation (GAmAge) is associated with maternal prenatal exposures and immediate birth outcomes. However, the association of GAmAge with long-term overweight or obesity (OWO) trajectories is yet to be determined.

**Methods:** GAmAge was calculated for 831 children from a US predominantly urban, low-income, multi-ethnic birth cohort using Illumina EPIC array and cord-blood DNA samples. Repeated anthropometric measurements aligned with pediatric primary care schedule allowed us to calculate body-mass-index percentiles (BMIPCT) at specific age and to define long-term weight trajectories from birth to 18 years.

**Results:** Four BMIPCT trajectory groups described the long-term weight trajectories: stable (consistent OWO: “early OWO”; constant normal weight: “NW”) or non-stable (OWO by year 1 of follow-up: “late OWO”; OWO by year 6 of follow-up: “NW to very late OWO”) BMIPCT. were used GAmAge was a predictor of long-term obesity, differentiating between group with consistently high BMIPCT and group with normal BMIPCT patterns and groups with late OWO development. Such differentiation can be observed in the age periods of birth to 1year, 3years, 6years, 10years, and 14years (p<0.05 for all; multivariate models adjusted for GEAA, maternal smoking, delivery method, and child’s sex). Birth weight was a mediator for the GAmAge effect on OWO status for specific groups at multiple age periods.

**Conclusions:** GAmAge is associated with BMI trajectories from birth to age 18 years, independent of GEAA and birth weight. If further confirmed, GAmAge may serve as an early biomarker for future OWO risk.

## Introduction

Biological aging, a process not completely correlated with chronological age, can be assessed using biological molecules and markers, (1–5), including DNA methylation (DNAm) (6–8). The biological age predicted by DNAm (mAge) has been studied for the correlation with chronological age and associations with health outcomes over the past decade; The mAge was found to be associated with body mass index (BMI) (9), abdominal adipose tissues (10), and liver fat (10). The residuals of mAge regressed of age (age acceleration), or the differences between methylation and chronological ages (“age diff” or Λage), is considered a strong predictor of all-cause mortality (11,12), cardiovascular mortality (13), and the incidence of cardiovascular disease (14).

Age prediction by DNAm and the associations with morbidity are not limited to adults. In newborns and children, epigenetic clocks can estimate the gestational and chronological age using DNA extracted from different tissues (saliva, peripheral, and cord blood) (15). Cord-blood age acceleration (the residuals of gestational age (GEAA) methylation age (GAmAge) regressed on GEAA) was associated with maternal exposures such as vitamin D supplementation during pregnancy in a sample of White, African American, and Hispanic mothers (16), pre-pregnancy BMI and smoking in a sample of British mothers (17,18), and gestational diabetes in Chinese mothers (19). In European decedents’ newborns, GAmAge and cord-blood age acceleration were associated with higher cord-blood vitamin B12 levels (20), delivery method (c-section) [15,17], and immediate birth outcomes of weight, length, and head circumference (17,21). Data on the long-term associations of GAmAge and age acceleration measured in cord blood and child’s phenotypes are sparse; Cord blood age acceleration was directly associated with a child’s weight and height up to 6 months and inversely associated with a child’s weight at 10 years of age (21).

In this analysis, we used data from a multi-ethnic mostly Black and Hispanic population birth cohort with extended time points for anthropometric measurements from birth to 21 years. We examined GAmAge as a predictor of childhood obesity and longitudinal trajectories from birth up to age 18 years, as reflected by distinct overweight or obesity (OWO) groups. We also examined whether birth weight can mediate these long-term associations. All of the above associations were examined beyond the impact of GEAA using appropriate statistical models to elucidate further the role of DNAm-based biological age as an indicator of health status.

## Methods

This study included 831 mother–newborns pairs from the Boston Birth Cohort (BBC; registered in ClinialTrial.gov NCT03228875), a US predominantly urban, low-income, Black and Hispanic population. The BBC was initiated in 1998 with rolling enrollment at the Boston Medical Center in Boston, MA, as detailed elsewhere (22,23). In brief, mothers who delivered a singleton live birth at the Boston Medical Center were invited to participate 24-72 hours after a vaginal delivery. The BBC is enriched by preterm (< 37 weeks of gestation) and low birth weight (< 2500 g) births by design of over-sampling PTB at enrollment. Pregnancies resulting from in vitro fertilization, multiple gestations (e.g., twins, triplets), fetal chromosomal abnormalities, major birth defects, or preterm birth due to maternal trauma were excluded. After mothers provided written informed consent, research assistants (RAs) administered a standardized questionnaire interview on maternal sociodemographic characteristics, lifestyle, including smoking and alcohol consumption, diet, and reproductive and medical history. Maternal and newborn clinical information, including birth outcomes, was abstracted from the medical records. The study protocol has received initial and annual approval from the Institutional Review Boards (IRBs) of Boston Medical Center and the Johns Hopkins Bloomberg School of Public Health.

### Main covariates

*Mother-child characteristics:* For background characteristics and adjustment of the statistical models, we used the following data: maternal age at delivery, parity (nulliparous or multiparous), maternal education (below college or college and higher), maternal self-reported race (Black/African American, White, and Hispanic), maternal pre-pregnancy BMI, maternal diabetes (non, gestational diabetes or pre-existing diabetes mellitus), delivery method, child’s sex (female versus male), maternal smoking, birth weight (as continuous and binary with above/below 2500g defined as the low birth weight (LBW) (24)), birth length, birth head circumference, and GEAA as continuous outcomes presented as weeks. We further characterized groups according to delivery week: extremely preterm (<28 weeks), very preterm (28 to 32 weeks), moderate to late preterm (week 32 to 37; WHO definitions), term (37 to 41 weeks), late-term (41 to 42 weeks), and post-term (>42 weeks; ACOG definitions; (25)). The estimation of GEAA was detailed before (22) and was performed using an established algorithm based on both the last menstrual period and the result of early ultrasound (<20 weeks’ gestation). Fetal growth groups– small for gestational age (SGA), appropriate for gestational age (AGA), and large for gestational age (LGA) were determined by the birth weight and gestational age as described before (26).

### Long-term obesity groups and BMIPCT

Out of the data available for the BBC, we selected 3029 children with height and weight measurements with sufficient follow-up data. We calculated BMIPCT using WHO (birth to 2 years old; (27)) and CDC growth charts (age 2 years and up;(28)) for these children. As child well-care visits had different frequencies, the BMIPCT data was divided into the following 32 time windows: monthly measurements in the first year, quarterly measurements in the second, and yearly measurements from month 36 (3^rd^ year) to the 216th month. Out of the 3029 children with BMIPCT, 940 had available DNAm measured.

*Obesity-related age periods*: In accordance with our previous work (29), we primarily focused on seven age periods representing different developmental phases previously identified as obesity-related critical periods: birth to 1 year old, birth to 2 years, birth to 3 years, birth to 6 years, birth to 10 years, birth to 14 years and birth to 18 years. Based on the first age period in this analysis (birth to 1) and the availability of children with DNAm analysis, we selected children with at least one BMIPCT measurement at each age period. We refined the similar sample size at each period, thus resulting in a sample size of 831 for each age period allowing us to follow the same sample of children across multiple age periods for their observed or discovered BMIPCT trajectory.

*BMIPCT missing data and OWO groups*: Imputation of BMIPCT missing data was detailed before (30) and in **Supplemental methods 1**. The OWO groups were constructed separately for each period, as follows: first, we applied k-means clustering with k=2. Next, we used Principal Component Analysis (PCA) to find the 1^st^ and 2^nd^ principal components. Since the first principal component primarily determines the k-means clustering, we divided the two groups above into four using the 2^nd^ principal component, as previously demonstrated (31). The groups resulting from this procedure represent four distinctive OWO trajectories, with two consistent-weight groups and two non-consistent weight-increase groups named retrospectively after examining the trajectories, as published before (29): 1. Early OWO: children with early onset OWO who demonstrated a consistent high BMIPCT from birth to the end of each age period; 2. Late OWO: late onset OWO children that were NW at birth but experienced a rapid weight increase in the first months of life to become OWO by year one; 3. NW to very late OWO: children distinguished from the late OWO by maintaining NW at early ages but becoming OWO by year six; 4. NW children consistently kept NW from birth to the end of each age period.

### DNA methylation profiling and calculation of gestational methylation age

The blood draw procedure and quality control (QC) steps were detailed before (29,32). In summary, the labor and delivery service’s trained nursing staff obtained cord blood after delivery. Genome-wide DNA profiling from 963 samples (plus 21 replicates) was performed using the MethylationEPIC BeadChip (850K). (33). Sample-level QC: We excluded 23 samples: 7 sex mixed-up samples, 2 samples with call rate <98% methylation sites, 12 samples with mean log2 intensity < 10, and 2 samples with logistic error. Probe-level QC: We performed the single-sample Noob (ssNoob) methods for background and dye bias correction (34). For a total of 865,859 CpG sites, we extracted beta values. No probes were removed to calculate GAmAge, in accordance with a previous publication (35), which presented a GAmAge prediction model based on the EPIC array. GAmAge, measured in days, was calculated for 831 children with available DNAm data from cord blood samples and calculated OWO trajectory groups using the “methylclock” R package (36).

### Statistical analysis

The primary aim of this study is to examine the association between GAmAge and BMIPCT trajectories across several age periods during childhood. Summary statistics were performed to compare newborns’ demographic and clinical characteristics across OWO groups using the chi-square test for categorical variables and ANOVA for continuous variables. ANOVA post hoc correction for multiple comparisons was performed using Bonferroni correction. Pearson correlation was used to examine the correlation between continuous variables. Multinomial regression was used to associate OWO groups with GAmAge, with adjustment for covariates that may affect birth weight and week and were associated with OWO groups in univariate analysis: gestational age, child’s sex, maternal smoking, and delivery method. Linear regression models were used to examine the association with continuous dependent outcomes. Mediation analysis using the “mediation” R package (37) was performed to examine the mediatory role of birth weight in the association between GAmAge and OWO groups. Since OWO trajectory is a four-factor variable, we used logistic regression to perform the mediation analysis with the early OWO as the reference group and performed 3 comparisons for the mediation (reference group vs. late OWO or NW to very late or NW) per age period. All statistical analyses were performed using R (version 4.1; R Foundation for Statistical Computing).

## Results

### Population characteristics

Maternal and child characteristics across OWO birth to 1y groups were presented in **Table 1**. Significant differences were observed in the children’s sex (p=0.031), with the smallest relative number of girls in the NW to very late OWO group and maternal smoking (p=0.003), with most mothers reporting ever smoking in the NW to very late OWO group. The early OWO group had the highest birth weight (p=1.2e-10 vs. late OWO, p=1.5e-4 vs. NW). The NW to very late OWO group had the lowest birth weight compared with the early OWO and NW (p<2.2e-16 for both) and were born in an earlier week compared with the other three groups (p=1.6e-14 vs. early OWO, p=8.1e-07 vs. late OWO, p=9.1e-12 vs. NW). The NW to very late OWO had the highest percentage of children born preterm and SGA.

**Table 1:**
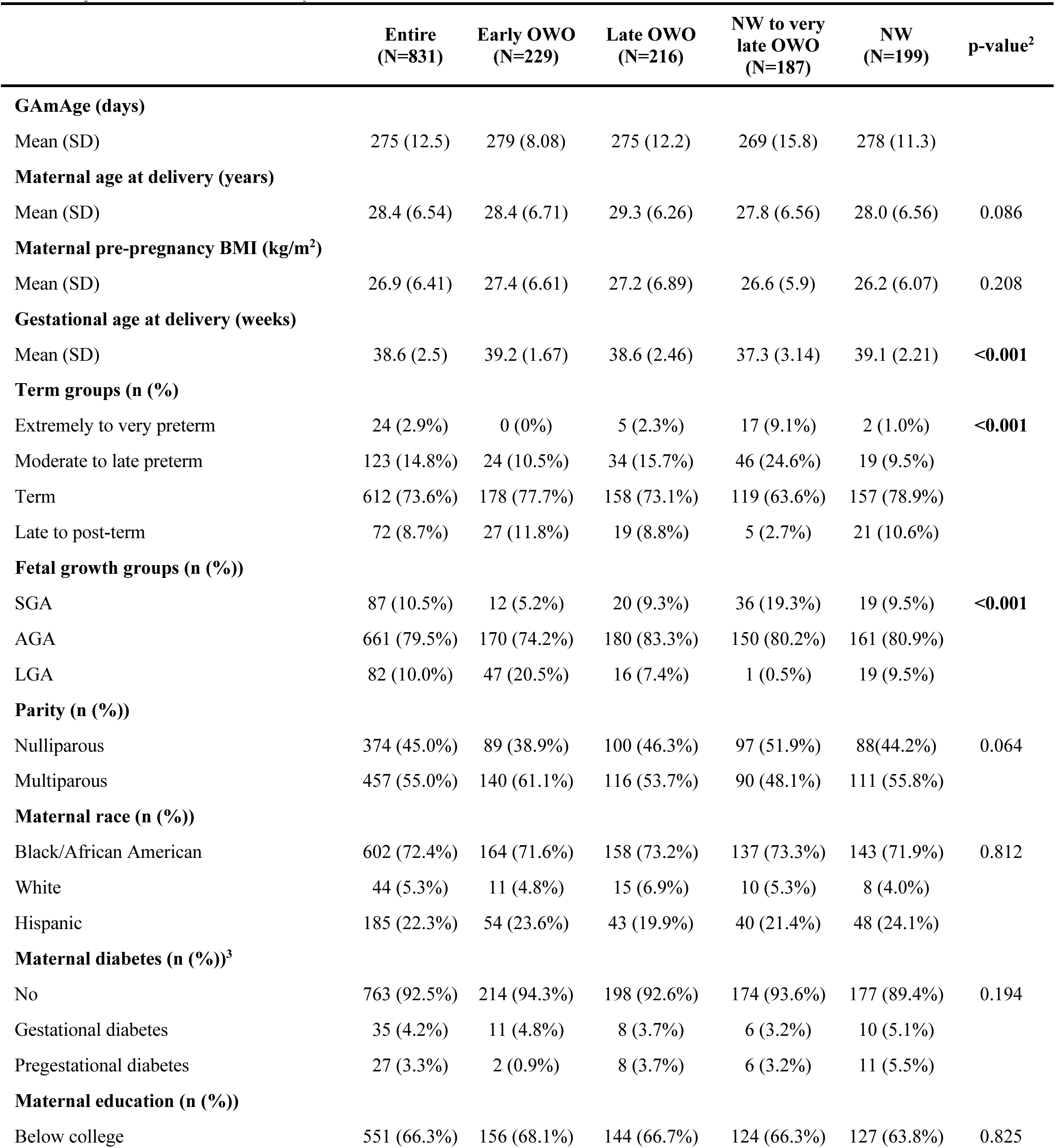
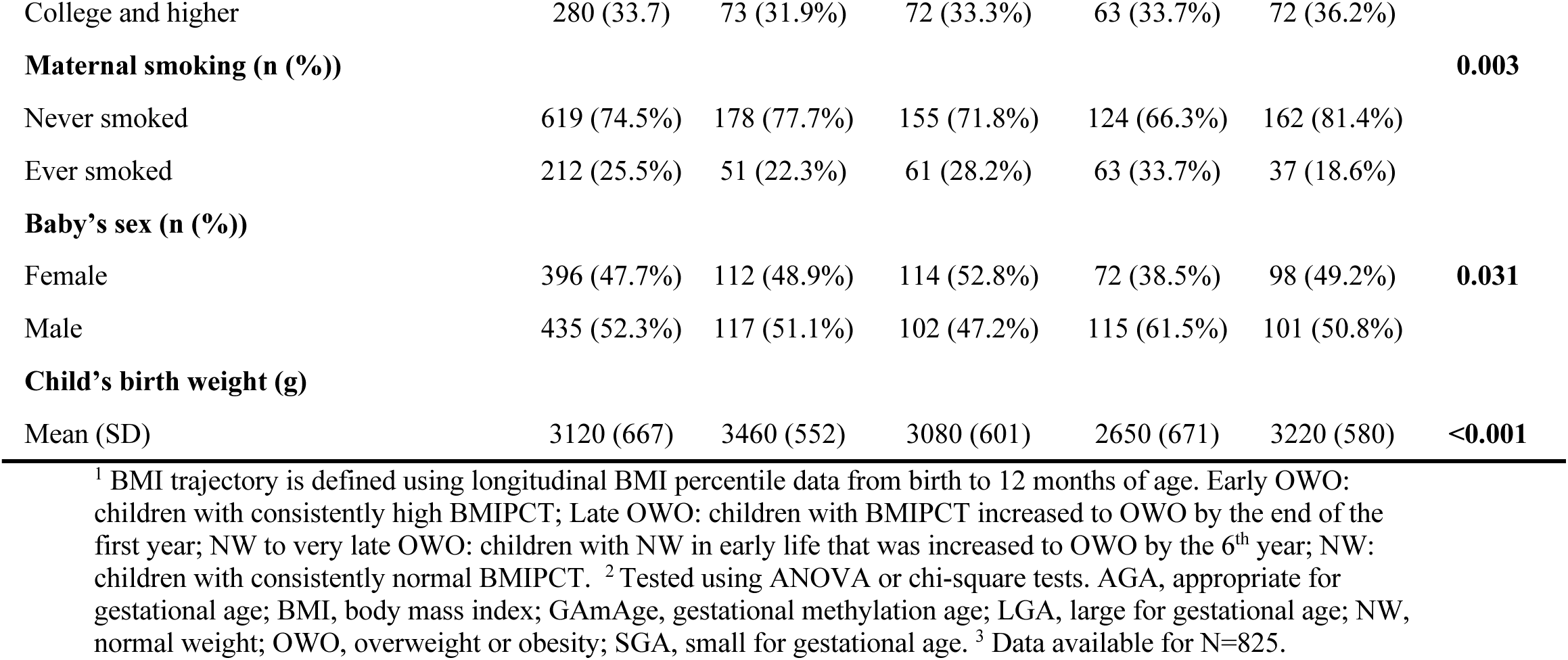
Prenatal and perinatal characteristics across subgroups of child BMI longitudinal trajectories from birth to 1y^1^.

### GAmAge associations with long-term obesity

GAmAge (275.5±12.5 days) and GEAA (270.0±17.5 days) were strongly correlated (r=0.89, p<2.2e-16). Stratifying by the OWO groups at each age period examined, we observed the strongest correlation of GAmAge and GEAA among NW to very late OWO children, compared with the other groups, across multiple age periods (**Figure 1; Figure S1**): early OWO vs. late OWO vs. NW to very late OWO vs. NW: 0.734 vs. 0.886 vs. 0.926 vs. 0.858, respectively (p<2.2e-16 for all).

**Figure 1:**
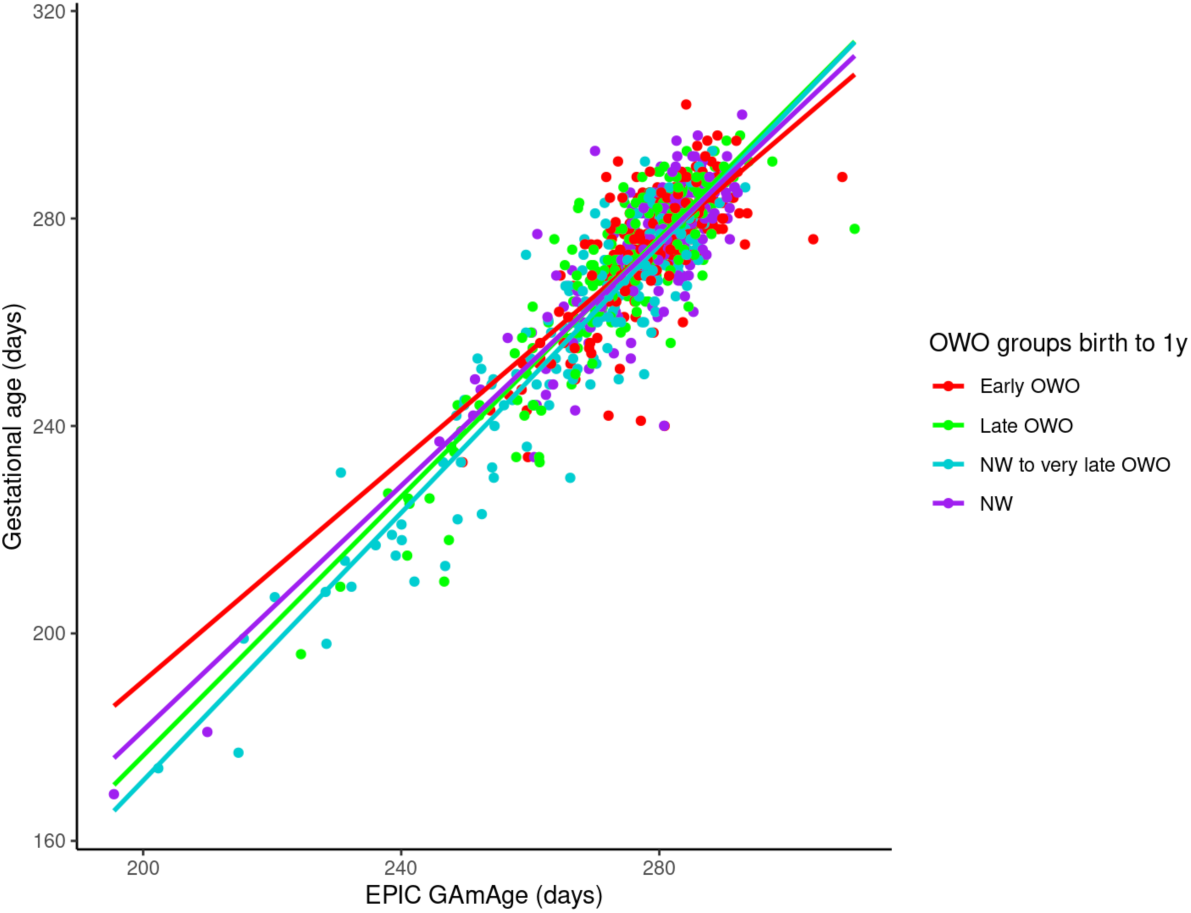
GAmAge and gestational age. The correlation between GAmAge and gestational age across OWO group at age period birth to 1y.

Next, we examined whether GAmAge could predict long-term BMIPCT patterns represented by OWO groups. The OWO groups differed in GAmAge in the following age periods (birth to 1: p=5.93e-16; birth to 2: 0.0279; birth to 3: p=0.0186; birth to 6: p=0.00865; birth to 10: p=0.00622; birth to 14: p=0.022; birth to 18: p=0.0478). Post-hoc correction for multiple comparisons showed that significant differences between early age periods (birth to 1, 2, 3, and 6 years) were mostly observed between the early OWO and the late OWO and NW to very late OWO (**Table S1**). For later age periods (birth to 10 and birth to 14), the differences were mostly between the early OWO and NW to very late OWO (p=0.00018 and p=0.001, respectively). After accounting for GEAA, child’s sex, delivery method, and maternal smoking, GAmAge was associated with OWO groups in multiple age periods: significantly higher GAmAge was observed in both consistent BMIPCT groups, early OWO and NW, compared with the non-consistent late and NW to very late OWO groups (**Figure 2**; **Table 2**). Setting NW as the reference group, the relative odds ratio of 0.98 for a one-unit increase in GAmAge in the NW vs. the very late OWO group was consistent for the age periods birth to 1 years, 3 years, and 6 years (p<0.05 for all). A similar observation for the relative odds ratio of 0.96-0.98 for a one-unit increase in GAmAge in the NW vs. the late OWO was found for age periods birth to 1 years, birth to 6 years, birth to 10 years, and birth to 14 years (p<0.05 for all).

**Figure 2:**
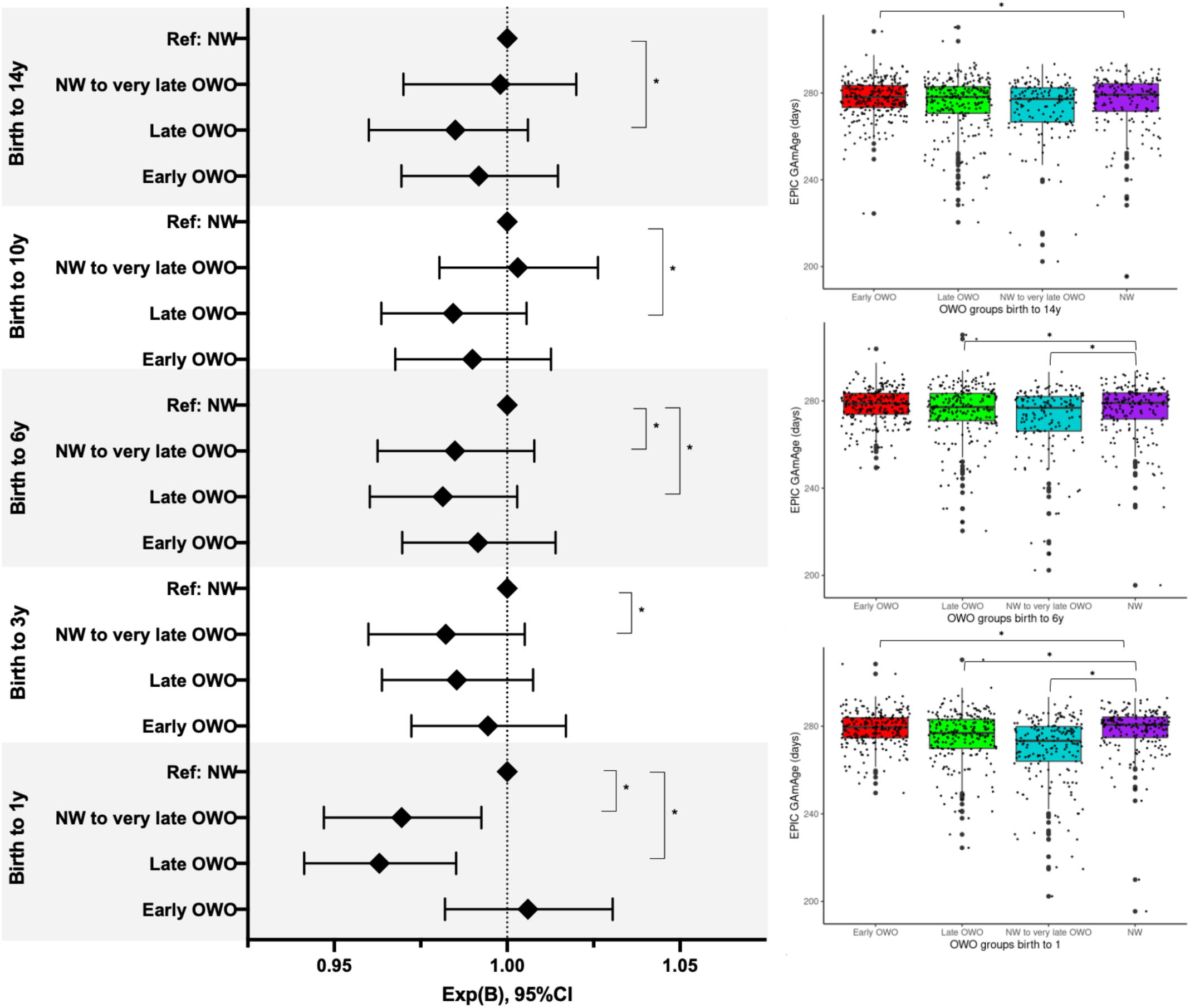
Child’s GAmAge across OWO groups in selected age periods. Left: results of the multinomial regression with NW as the reference group. Models adjusted for gestational age in days, maternal smoking, delivery method, and child sex. Results presented for the GAmAge and gestational age predictors. N=831. Right: box plots for GAmAge across OWO groups for age periods birth to 1, birth to 6, and birth to 18. GAmAge, gestational methylation age; NW, normal weight; OWO, overweight or obese. * Denotes significant difference at p<0.05 level.

**Table 2:**
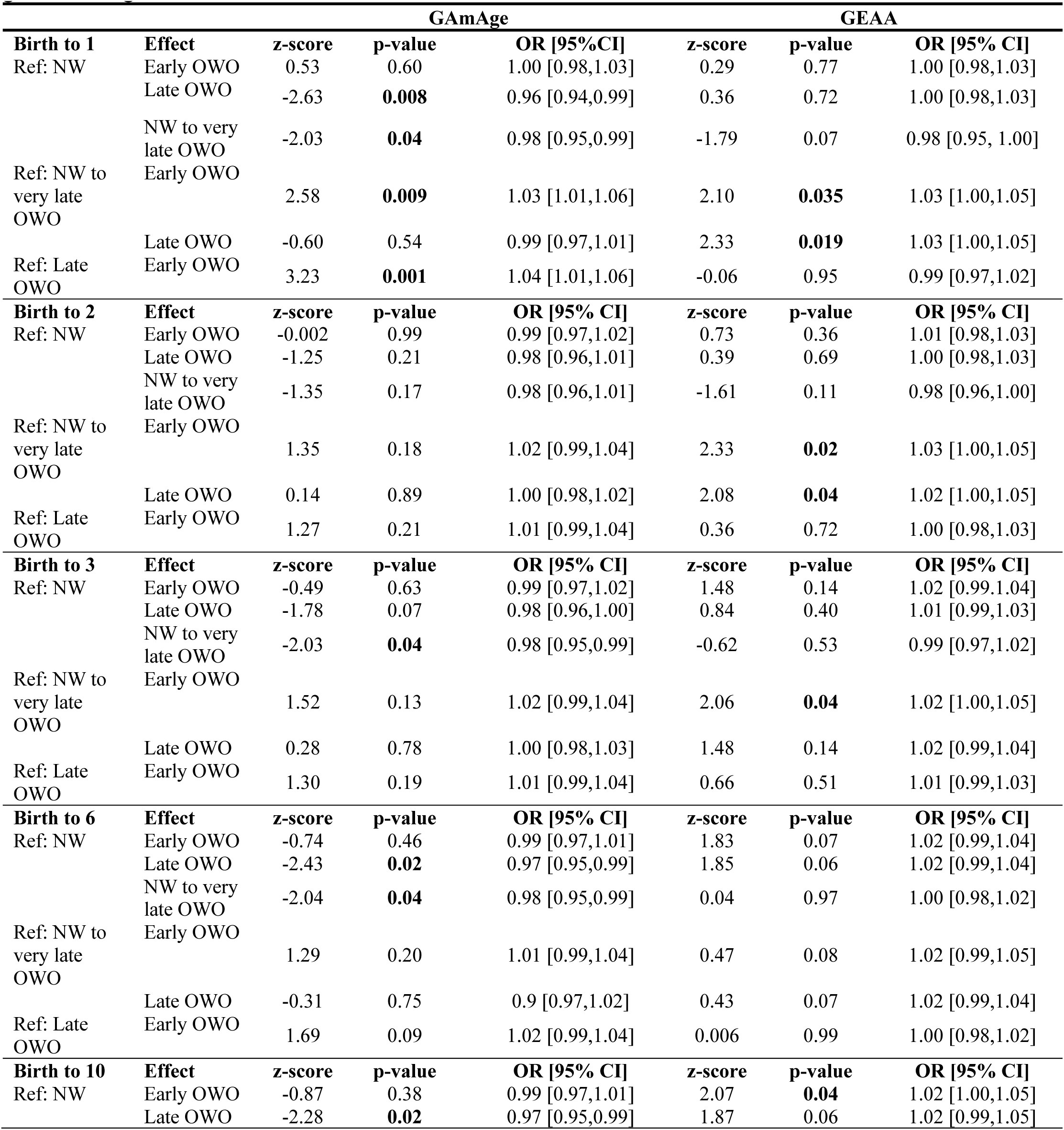
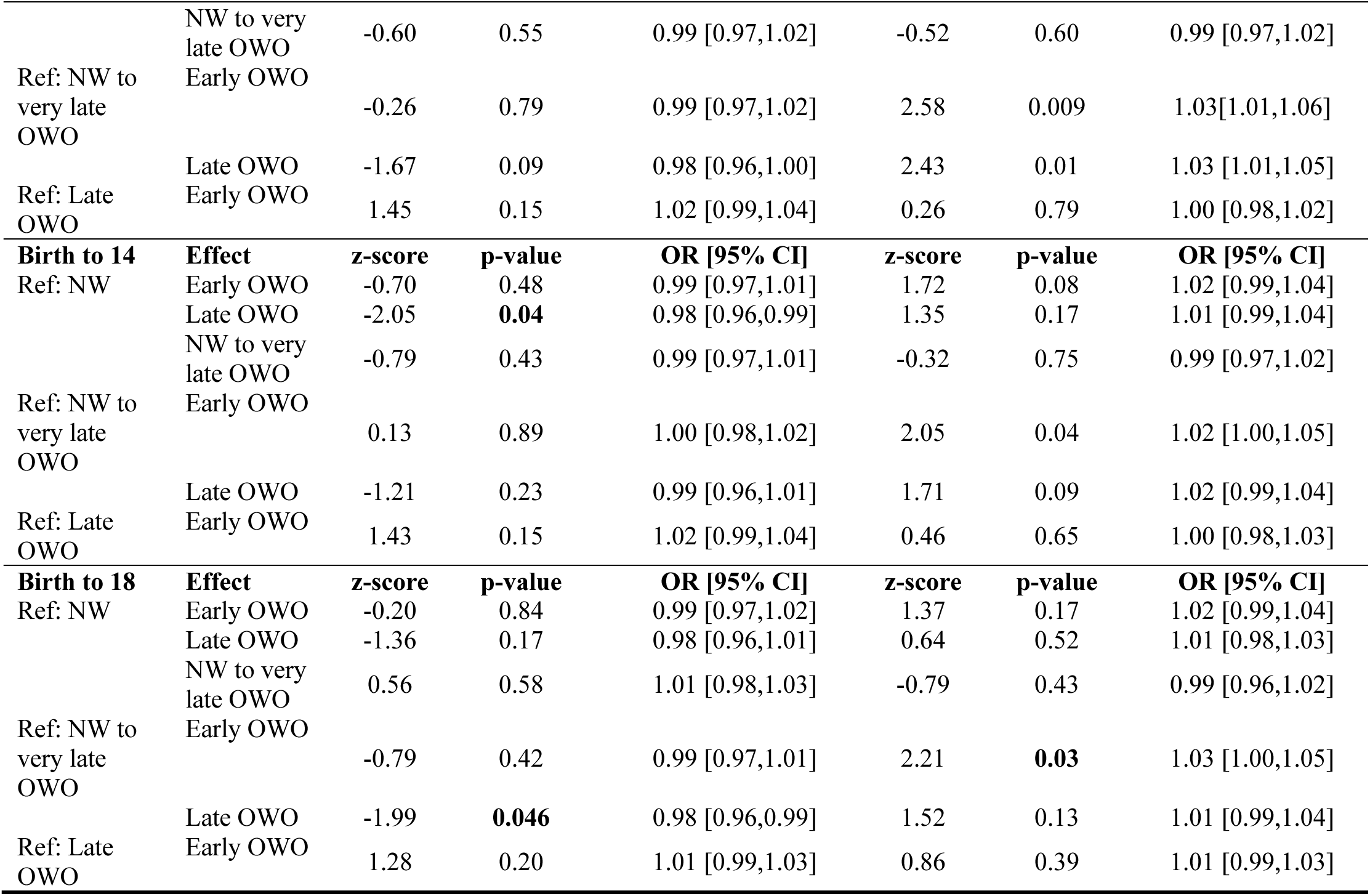
The association between GAmAge, GEAA, and OWO groups across different age periods. The model includes mutual adjustment for GAmAge and GEAA and the following covariates: maternal smoking, delivery method, and child sex. Results presented for the GAmAge and gestational age predictors. N=831. GAmAge, gestational methylation age; GEAA, gestational age

We also examined whether GAmAge provides additional information over GEAA to predict long-term OWO. We compared the GAmAge and GEAA coefficients in the multivariate models that included mutual adjustment for both predictors across several age periods (**Table 2**). In the age periods of birth to 1, 3, 6, 10, 14, and 18 years, GAmAge explained more than GEAA the differences between some OWO groups, as reflected by significant z-score (e.g., birth to 1, late OWO vs. early OWO: 3.23 vs. –0.06 z-scores for GAmAge vs. GEAA, respectively; birth to 3, NW vs. NW to very late OWO: –2.03 vs. –0.62; birth to 6, NW vs. late OWO: –2.43 vs. 1.85). On the other hand, at some age periods (birth to 1, 2, 3, 10, 14, and 18 years), GEAA explained more of the association with some OWO groups (e.g., birth to 1, late OWO vs. NW to very late OWO: –0.60 vs. 2.33 z-scores for GAmAge vs. GEAA, respectively; birth to 2, early OWO vs. NW to very late OWO: 1.35 vs. 2.33).

We repeated the analysis for the associations with long-term obesity to examine the association with GAmAge acceleration (the residuals from linear regression using GEAA as a predictor for GAmAge). Results from this analysis presented at **Table S2**. Unlike the associations in the joint model, using GAmAge acceleration instead of GAmAge and GEAA did not results in any significant associations with the OWO groups, in all age periods.

### GAmAge and GEAA contribution to birth weight variation

Birth weight was associated with GAmAge after adjusting for GEAA, child’s sex, delivery method, and maternal smoking (beta=7.66, p=0.0064). Examining the R2 of this model, i.e., how much variation in birth weight was explained by the model, starting with the association of birth weight with the child’s sex, delivery method, and maternal smoking (R2=0.031), adding GAmAge (R2=0.449) or GEAA (R2=0.516), showed the highest R2 with both GAmAge and GEAA in the model (R2=0.521), suggesting GAmAge explained additional 0.4% variation in birth weight on top of GEAA, sex, delivery method, and maternal smoking.

Since GEAA showed a similar high proportion of the explained variance in birth weight for the above model, we further stratified the birthweight model by subgroups of the delivery week. We found that the association of birth weight and GAmAge was the strongest in the extremely to very preterm strata (extremely preterm to very preterm: beta=21.60, p=0.024, R2=0.582 when both GAmAge and GEAA in the model; moderate to late preterm: beta=12.63, p=0.029, R2=0.305; term: beta=6.39, p=0.08, R2=0.198; late to post-term: beta=-11.13, p=0.264, R2=0.053). In the extremely to very preterm strata, for a model adjusted just for child’s sex, delivery methods, and maternal smoking (R2=0.059), adding GAmAge (R2=0.575) contributed more to the model than adding GEAA (R2=0.442). This was not observed within the moderate to late preterm strata, where adding GEAA to the model contributed more than adding GAmAge (R2 of a model without GAmAge or GEAA=0.048; R2 for adding GAmAge=0.221; R2 for adding GEAA instead of GAmAge=0.276). This was also observed within the term strata (R2=0.045, R2=0.1234, R2=0.194; for models without GAmAge or GEAA, a model with GAmAge added, a model with GEAA added, respectively).

### Mediation of the GAmAge association with OWO groups by birth weight

In the subsequent analysis, we examined whether birth weight mediated the association between GAmAge and long-term OWO groups. We performed a mediation analysis using the early OWO as a reference in a two-group comparison logistic model. We found that birth weight mediated the association between GAmAge and the OWO groups, consistently between the early OWO and the NW in the age periods of birth to 2, 6 years, 10 years, 14 years, and 18 years. A summary of the casual mediation analysis is presented in **Table S3**.

## Discussion

In our study of 831 children, the GAmAge was associated with long-term obesity and was lower in late and very late OWO trajectories compared with early OWO. These associations were mediated by birth weight in multiple age periods, specifically for the associations between the consistent BMIPCT groups: early OWO and NW.

We found that among children assigned to the group NW to very late OWO, the strongest correlation between GAmAge and GEAA was observed compared with the other OWO group at multiple age periods. The NW to very late OWO group was characterized by having the lowest birth week and the highest percentage of preterm and SGA children. This may reflect the differences in DNAm between preterm and term babies (38), as a previous study of 36 sex-matched preterm infants (birth < 33 weeks gestation) and 36 sex-matched term babies (birth > 37 weeks gestation) found 83 CpGs differentially methylated between the groups. Another study investigated the epigenetic impact of preterm birth in isolated hematopoietic cell populations (39). They concluded that some epigenetic markers in hematopoietic cells might also differ by prematurity due to differences in the preterm immune system compared with term neonates in both cell composition and function. These observations highlight the need to further investigate DNAm patterns and regulatory mechanisms among groups of delivery week.

The data on the associations between GAmAge and long-term weight trajectories are limited. In a prospective study that followed 785 children from birth to 10 years old (21), the association of regressed GAmAge of GEAA was directly associated with the increase in age-specific time windows weight measurements up to 6 months. However, these associations reversed from the age of 5 years onwards, and the regressed GAmAge of age was inversely associated with the child’s weight: a non-significant trend in the ages of 5 to 9 years and a significant association at the age of 10 were observed. In our analysis, we demonstrated the associations of GAmAge with OWO patterns in several age periods from birth to 18 years. The use of epigenetic markers as an early indicator for later life obesity was also demonstrated in our previous epigenome-wide association study, where specific DNAm sites were associated with OWO trajectory patterns, differentiating between the OWO groups (29). Here, we found that consistent BMIPCT trajectories groups early OWO (children with elevated BMIPCT from birth) and NW (children with NW pattern from birth) significantly differ in GAmAge from the non-consistent BMIPCT trajectories groups late (OWO by the end of year 1) and very late OWO (NW until 6 years old, and OWO onwards). The accumulating evidence for the predictive ability of early-life epigenetic signatures on later-life obesity should be further examined. Moreover, the associations of early epigenetic signatures with later-life morbidity should also be studied.

Birth weight was a mediator for this association up to year 2, and an independent predictor for OWO trajectories in later age periods. The two non-consistent BMIPCT trajectory groups started with a median BMIPCT below the 50th percentile, but by the end of year 1 and year 6, respectively, children in these groups become OWO. Birth weight has been studied for the associations with short– and long-term obesity and other health outcomes; a U-shaped association between birth weight and childhood obesity was observed in a cohort of 5141 children between the ages of 9 to 11 (40). In this study, beyond factors such as highest parental education, maternal history of gestational diabetes, child age, infant feeding mode, gestational age, unhealthy diet pattern scores, and sleep quality, the odds ratio of being >4000g at birth was 1.77 for boys and 2.48 for girls. Also, children from high-income countries had a higher risk of childhood obesity with birth weight > 4000g, whereas children from low– or middle-income countries had an increased risk starting at 3500g of birth weight. On the other hand, low birth weight was associated with cardiometabolic diseases in adulthood (41) and with childhood and adulthood obesity (42,43). Yet, it has to be noted that not all findings indicate that low birth weight might lead to childhood obesity (42). Therefore, utilizing GAmAge as an independent marker at birth may assist in identifying late-onset obesity in children that are NW and with a lower birth week in their early life without other indication for the long-term OWO trajectory.

There are some limitations to this study. In the casual mediation analysis – birth weight and GAmAge were measured at the same time. Second, the findings’ reproducibility depends on available birth cohorts with dense repeated BMIPCT measurements and DNAm, as the BBC has. The strengths of this study, beyond its large sample size and extended time points for BMIPCT measurements from birth to 18 years, are the novel associations described of GAmAge with long-term OWO trajectories.

In conclusion, biological signatures based on DNAm are independent of GEAA in long-term association with OWO. GAmAge may early-detect the onset of late and very late OWO.

## Data availability statement

The data, data dictionary, and analytical programs for this manuscript are not currently available to the public. However, they can be made available upon reasonable request and after the review and approval of the institutional review board.

## Authors’ contribution

Anat Yaskolka Meir, Xiumei Hong, Xiaobin Wang, and Liming Liang were responsible for study conception. Xiaobin Wang supervised collection of phenotypic data and biospecimens. Anat Yaskolka Meir and Liming Liang verified the underlying data. Anat Yaskolka Meir was responsible for drafting of the manuscript. Xiumei Hong, Guoying Wang, Liming Liang, and Xiaboin Wang supervised DNA methylation data generation. Anat Yaskolka Meir and Xiumei Hong performed DNA methylation quality control and data cleaning. Anat Yaskolka Meir performed most of the statistical analyses under the guidance and technical support of Liming Liang. Xiaboin Wang was responsible for overseeing acquisition of the epidemiological and clinical data as well as biospecimens. All the authors were responsible for critical review and revision of the manuscript and contributed to data interpretations. All authors read and approved the final version of the manuscript.

## Ethics

Written informed consent was obtained from all the study mothers. Institutional Review Boards of the Boston Medical Center and the Johns Hopkins Bloomberg School of Public Health approved the study. The study is also registered on ClinicalTrials.gov (NCT03228875).

