## Supplemental material for "Newborn DNA methylation age differentiates long-term weight trajectory: The Boston Birth Cohort"

**Supplemental methods 1:** BMIPCT imputation

To complete missing data (for the primary analysis), we fitted a LOWESS (locally weighted scatterplot smoothing) curve [44], as we demonstrated. Briefly, we adopted a locally weighted scatterplot smoothing curve within a time-limit aware scheme (LOWESS-TLS). Each missing BMIPCT data was predicted using a regression curve (LOWESS) of the available observations within the reasonable time frame around the missing time point.

### Table S1: Differential level of GAmAge across OWO groups in different age periods

| Age period | ANOVA P-value | P-value for Bonferroni correction | | | | | |
|---|---|---|---|---|---|---|---|
| | | Early OWO vs. late OWO | Early OWO vs. very late OWO | Early OWO vs. NW | Late OWO vs. NW to very late OWO | Late OWO vs. NW | Very late OWO vs. NW |
| **Birth to 1y** | 5.93e-16 | 0.0038 | 1.4e-14 | 1 | 1.7e-05 | 0.0492 | 9.4e-12 |
| **Birth to 2y** | 0.0279 | 0.28 | 6.6e-11 | 1 | 4.6e-06 | 1 | 1.7e-08 |
| **Birth to 3y** | 0.0186 | 0.08834 | 4.8e-09 | 1 | 0.00083 | 1 | 6.6e-06 |
| **Birth to 6y** | 0.00865 | 0.1588 | 2.2e-06 | 0.5391 | 0.0117 | 1 | 0.0048 |
| **Birth to 10y** | 0.00622 | 0.13499 | 0.00018 | 0.29081 | 0.18584 | 1 | 0.20816 |
| **Birth to 14y** | 0.022 | 0.085 | 0.001 | 0.674 | 0.584 | 1 | 0.252 |
| **Birth to 18y** | 0.0478 | 0.010 | 0.071 | 0.452 | 1 | 1 | 1 |

**Table S2: The association between age acceleration and OWO groups across different age periods**. Model adjusted for maternal smoking, delivery method, and child sex. N=831. GAmAge, gestational methylation age.

| | | GAmAge acceleration | | |
|---|---|---|---|---|
| **Birth to 1** | **Effect** | **z-score** | **p-value** | **OR [95%CI]** |
| Ref: NW | Early OWO | 0.24 | 0.81 | 1.00 [0.97,1.04] |
| | Late OWO | -1.48 | 0.13 | 0.97 [0.94,1.01] |
| | NW to very late OWO | -1.27 | 0.20 | 0.98 [0.94,1.01] |
| Ref: NW to very late OWO | Early OWO | 1.55 | 0.89 | 1.03 [0.99,1.06] |
| | Late OWO | -0.14 | 0.12 | 0.99 [0.96,1.03] |
| Ref: Late OWO | Early OWO | 1.79 | 0.07 | 1.03 [0.99,1.06] |
| **Birth to 2** | **Effect** | **z-score** | **p-value** | **OR [95% CI]** |
| Ref: NW | Early OWO | -0.15 | 0.88 | 0.99 [0.97,1.03] |
| | Late OWO | -0.75 | 0.45 | 0.99 [0.95,1.02] |
| | NW to very late OWO | -0.98 | 0.33 | 0.98 [0.95,1.02] |
| Ref: NW to very late OWO | Early OWO | 0.87 | 0.38 | 1.01 [0.98,1.05] |
| | Late OWO | 0.28 | 0.77 | 1.00 [0.97,1.04] |
| Ref: Late OWO | Early OWO | 0.63 | 0.53 | 1.01 [0.82,1.79] |
| **Birth to 3** | **Effect** | **z-score** | **p-value** | **OR [95% CI]** |
| Ref: NW | Early OWO | -0.55 | 0.58 | 0.99 [0.96,1.02] |
| | Late OWO | -1.13 | 0.26 | 0.98 [0.95,1.01] |
| | NW to very late OWO | -1.38 | 0.16 | 0.97 [0.94,1.01] |
| Ref: NW to very late OWO | Early OWO | 0.93 | 0.35 | 1.02 [0.98,1.05] |
| | Late OWO | 0.40 | 0.73 | 1.00 [0.97,1.04] |
| Ref: Late OWO | Early OWO | 0.63 | 0.53 | 1.01 [0.98,1.04] |
| **Birth to 6** | **Effect** | **z-score** | **p-value** | **OR [95% CI]** |
| Ref: NW | Early OWO | -0.71 | 0.47 | 0.99 [0.96,1.02] |
| | Late OWO | -1.60 | 0.11 | 0.97 [0.94,1.01] |
| | NW to very late OWO | -1.39 | 0.17 | 0.97 [0.94,1.01] |
| Ref: NW to very late OWO | Early OWO | 0.79 | 0.43 | 1.01 [0.98, 1.05] |
| | Late OWO | -0.07 | 0.95 | 0.99 [0.96,1.03] |
| Ref: Late OWO | Early OWO | 0.96 | 0.34 | 1.02 [0.98,1.05] |
| **Birth to 10** | **Effect** | **z-score** | **p-value** | **OR [95% CI]** |
| Ref: NW | Early OWO | -0.76 | 0.44 | 0.99 [0.95,1.02] |
| | Late OWO | -1.49 | 0.14 | 0.97 [0.94,1.01] |
| | NW to very late OWO | -0.45 | 0.65 | 0.99 [0.95,1.03] |
| Ref: NW to very late OWO | Early OWO | -0.26 | 0.79 | 0.99 [0.96,1.03] |
| | Late OWO | -0.97 | 0.33 | 0.98 [0.95,1.02] |
| Ref: Late OWO | Early OWO | 0.82 | 0.41 | 1.01 [0.98,1.04] |
| **Birth to 14** | **Effect** | **z-score** | **p-value** | **OR [95% CI]** |
| Ref: NW | Early OWO | -0.63 | 0.53 | 0.99 [0.96,1.02] |
| | Late OWO | -1.33 | 0.18 | 0.98 [0.94,1.01] |
| | NW to very late OWO | -0.56 | 0.57 | 0.99 [0.95,1.03] |

| | | | | |
|---|---|---|---|---|
| Ref: NW to very late OWO | Early OWO | 0.02 | 0.0.99 | 1.00 [0.97,1.04] |
| | Late OWO | -0.67 | 0.50 | 0.99 [0.95,1.02] |
| Ref: Late OWO | Early OWO | 0.44 | 0.42 | 1.01 [0.98,1.04] |
| **Birth to 18** | **Effect** | **z-score** | **p-value** | **OR [95% CI]** |
| Ref: NW | Early OWO | -0.27 | 0.79 | 0.99 [0.96,1,03] |
| | Late OWO | -0.89 | 0.37 | 0.98 [0.95,1.02] |
| | NW to very late OWO | 0.35 | 0.73 | 1.01 [0.97,1.05] |
| Ref: NW to very late OWO | Early OWO | -0.66 | 0.51 | 0.99 [0.95,1.02] |
| | Late OWO | -1.26 | 0.21 | 0.98 [0.94,1.01] |
| Ref: Late OWO | Early OWO | 0.73 | 0.46 | 1.01 [0.98,1.04] |

### Table S3: Mediation analysis

| **Age period** | **Ref + OWO group** | **p-value: the effect of GAmAge on the mediator (A)** | ***Average causal mediation effect (B)*** | **p-value** |
|---|---|---|---|---|
| **Birth to 1y** | Early OWO vs. late OWO | 0.195 | Na | Na |
| | Early OWO vs. very late OWO | 0.00556 | -2.73e-07 | **0.012** |
| | Early OWO vs. NW | 0.035 | -5.75e-04 | 0.068 |
| **Birth to 2y** | Early OWO vs. late OWO | 0.087 | Na | Na |
| | Early OWO vs. very late OWO | 0.087 | Na | Na |
| | Early OWO vs. NW | 0.0103 | -3.06e-03 | **0.024** |
| **Birth to 3y** | Early OWO vs. late OWO | 0.0468 | -7.73e-05 | 0.14 |
| | Early OWO vs. very late OWO | 0.039 | -1.11e-04 | 0.072 |
| | Early OWO vs. NW | 0.02115 | -1.72e-03 | 0.05 |
| **Birth to 6y** | Early OWO vs. late OWO | 0.04 | -2.83e-05 | 0.12 |
| | Early OWO vs. very late OWO | 0.04 | -6.04e-05 | 0.056 |
| | Early OWO vs. NW | 0.031 | -0.0007 | 0.05 |
| **Birth to 10y** | Early OWO vs. late OWO | 0.033 | -4.4e-05 | 0.098 |
| | Early OWO vs. very late OWO | 0.137 | Na | Na |
| | Early OWO vs. NW | 0.029 | -1.38e-04 | 0.056 |
| **Birth to 14y** | Early OWO vs. late OWO | 0.0236 | -7.11e-05 | 0.068 |
| | Early OWO vs. very late OWO | 0.006 | -0.003 | **0.01** |
| | Early OWO vs. NW | 0.014 | -2.64e-04 | **0.048** |
| **Birth to 18y** | Early OWO vs. late OWO | 0.0259 | -0.000133 | 0.08 |
| | Early OWO vs. very late OWO | 0.040 | -0.0006 | 0.086 |
| | Early OWO vs. NW | 0.015 | -8.61e-04 | **0.028** |

(A) Linear regression model: birth weight ~ GAmAge + gestational age + child's sex + delivery method + maternal smoking. (B) Causal mediation analysis using the following regression models: linear model (A) and a logistic regression model to assess the effect of the mediator birth weight on the two OWO groups: Ref + OWO group ~ GAmAge + birth weight + gestational age + child's sex + delivery method + maternal smoking. With GAmAge as the treat and birth weight as the mediator.

**Figure S1**: GAmAge and GEAA correlations across OWO groups in different age periods. a. Birth to 2 y. Correlation for early OWO vs. late OWO vs. NW to very late OWO vs. NW: 0.766 vs. 0.868 vs. 0.937 vs. 0.857, respectively (p<2.2e-16 for all). b. Birth to 3 y. Correlation for early OWO vs. late OWO vs. NW to very late OWO vs. NW: 0.749 vs. 0.877 vs. 0.938 vs. 0.868, respectively (p<2.2e-16 for all). c. Birth to 6 y. Correlation for early OWO vs. late OWO vs. NW to very late OWO vs. NW: 0.764 vs. 0.880 vs. 0.931 vs. 0.888, respectively (p<2.2e-16 for all). d. Birth to 10 y. Correlation for early OWO vs. late OWO vs. NW to very late OWO vs. NW: 0.790 vs. 0.870 vs. 0.935 vs. 0.894, respectively (p<2.2e-16 for all). e. Birth to 14 y. Correlation for early OWO vs. late OWO vs. NW to very late OWO vs. NW: 0.795 vs. 0.886 vs. 0.919 vs. 0.907, respectively (p<2.2e-16 for all). f. Birth to 18 y. Correlation for early OWO vs. late OWO vs. NW to very late OWO vs. NW: 0.818 vs. 0.899 vs. 0.902 vs. 0.902, respectively (p<2.2e-16 for all).

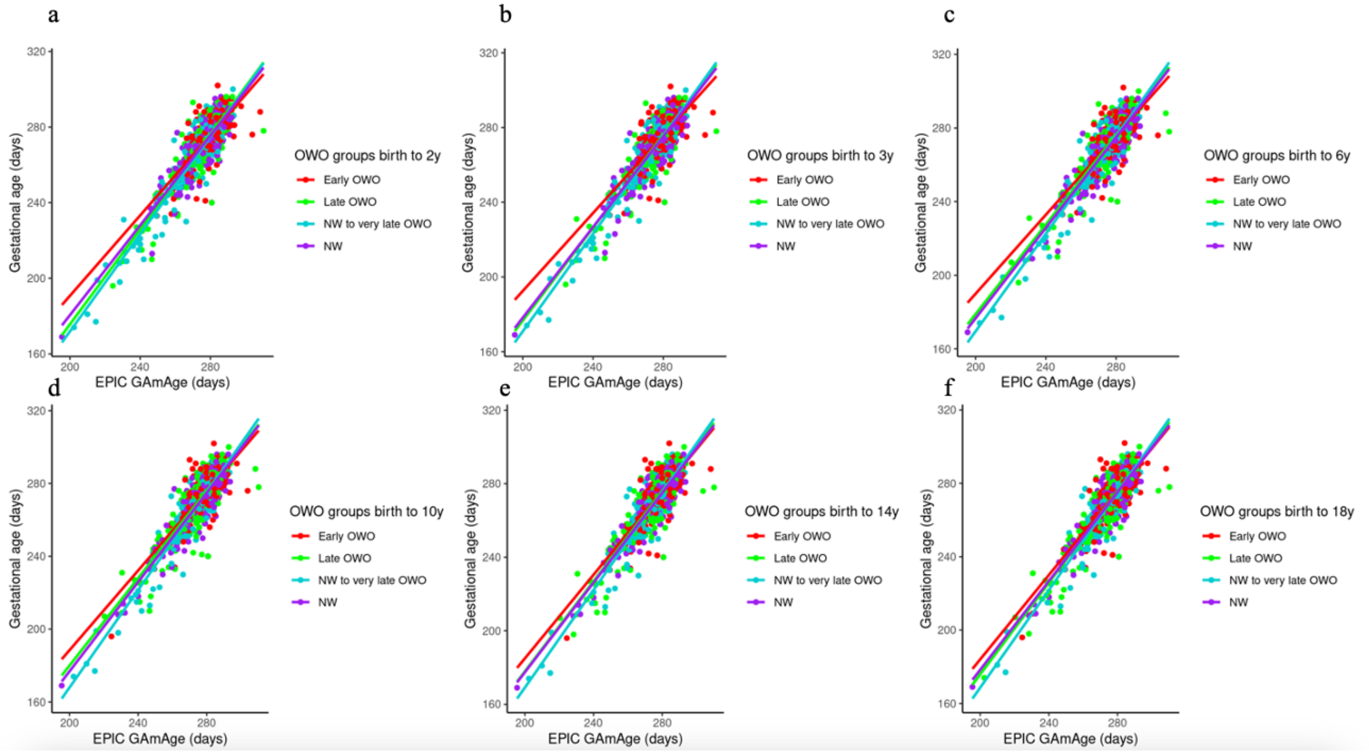
